# Fertility Transition in India, 1985-2020: Selection of the Aggregation Function Matters

**DOI:** 10.1101/2024.08.02.24310797

**Authors:** Aalok Ranjan Chaurasia

## Abstract

This paper analyses fertility transition in India during 1985-2020 based on the data from the official sample registration system. The analysis reveals that fertility transition in the country is contingent upon the way age-specific fertility rates are aggregated into a single composite indicator of fertility. When the simple arithmetic mean of age-specific fertility rates is used as a composite indicator of fertility, fertility in India has decreased almost linearly. However, when the geometric mean of age-specific fertility rates is used as the composite indicator of fertility, fertility transition in India appears to have stalled the period 2011-2013. The analysis also reveals that the change in marital fertility accounted for only about 35 per cent of the change in the simple arithmetic mean of age-specific fertility rates but more than half of the change in the geometric mean of age-specific fertility rates. The paper suggests that fertility transition should not be analysed in terms of the trend in the simple arithmetic mean of age-specific fertility rates or, equivalently, total fertility rate but should be analysed in terms of the trend in the geometric mean of age-specific fertility rates.

## Background

The total fertility rate (TFR) in India has now decreased to below replacement level (Government of India, 2022a; Government of India, 2022b; United Nations, 2022). According to the official sample registration system of the country, the TFR in the country decreased from 4.3 births per woman of childbearing age in 1985 to 2 births per women of childbearing age in 2020 (Government of India, 2022a). The latest (2019-2021) round of the National Family Health Survey (NFHS) has also estimated a TFR of about 1.99 births per woman of childbearing age during the period 2017-2019 (Government of India, 2022b) compared to a TFR of 3.4 births per woman of childbearing age during the period 1990-1992 (Government of India, 1995). The National Population Policy 2000 of India had targeted to achieve the replacement fertility (TFR=2.1) by the year 2010 (Government of India, 2000). This goal could be achieved only after a lag of almost 10 years. The delay in achieving the replacement fertility has implications for population stabilisation in India. The National Population Policy 2000 had projected that population of the country would stabilise by the year 2045 under the assumption that the replacement fertility would be achieved by the year 2010. The latest population projections prepared by the United Nations suggest that it is the most likely that the population of the country will continue to increase at least up to 1960 (United Nations, 2024).

The decrease in the TFR to the replacement or below replacement level has raised interest of demographers in the analysis of fertility transition in India. A recent study has concluded that India has followed an alternative pathway to low TFR which is different from the pathway followed by high-income countries (Park et al, 2023). Indian women continue to marry and produce births at young ages while the age at last birth has decreased so that births have increasingly got concentrated in the younger ages of the childbearing period. An increasing proportion of married Indian women go for sterilisation following the birth of two children to stop childbearing so that average fertility has decreased because of termination of childbearing at a young age (Park et al, 2023). The study has, however, not discussed the implications of fertility transition pathway followed by India for population stabilisation which is the medium-term goal of the National Population Policy 2000 (Government of India, 2000). It is well-known that population continues to increase for some time even after the achievement of replacement fertility because of the momentum of growth built in population age structure (Frejka, 1982; Keyfitz, 1971; Merrick, 1989). The impact of momentum on future population growth cannot be eliminated as it is the result of past trend in fertility and mortality but can be delayed through the increase in the mean age at childbearing with the decrease in fertility through increasing the age at first birth and the period between successive births (Bongaarts, 1994).

Achievement of below replacement fertility in India also masks variation in TFR within the country, across states and Union Territories. The sample registration system provides estimates of TFR for 22 states of the country and, in 2020, the TFR ranged from 1.4 births per woman of childbearing age in the National Capital Territory of Delhi, Tamil Nadu and West Bengal to 3 births per woman of childbearing age in Bihar in these states. The latest (2019-2021) round of NFHS provides estimates of TFR for all 36 states/Union Territories of the country and informs that TFR ranged from 1.05 births per woman of childbearing age in Sikkim to 2.98 births per woman of childbearing age in Bihar during the three years prior to the survey (2017-2019). There are five states in the country where TFR remains above the replacement level. These include Uttar Pradesh, the most populous state of the country and Bihar (Government of India, 2022b). District level estimates of TFR are not available from either the sample registration system or the NFHS. In the past, district level estimates of TFR were prepared using data from decennial population censuses (Government of India, 1988; 1997; Guilmoto and Rajan, 2002; 2013; Mishra et al, 1994) but there has been no population census in the country after 2011. An indirect approach, using data from the latest round of NFHS, however, suggests that TFR was below replacement level in only 326 of the 707 districts of the country.

Fertility transition encompasses change in the fertility experience of women of childbearing age and the change in the age pattern of fertility. At the aggregate level, the change in the fertility experience of women of childbearing age is universally captured through the change in TFR which is the sum of the age-specific fertility rates. The change in TFR does not reflect the change in the age pattern of fertility. TFR is sensitive to the shift in the age pattern of fertility or the timing of fertility (Hajnal, 1947). The postponement of births leads to an increase in the mean age at childbearing which affects age-specific fertility rates and hence TFR (Hajnal, 1947; Feeney, 1983; Ryder, 1964). The change in TFR, therefore, may be affected by the change in the timing of births or the tempo effect and the number of births women have when they end childbearing or the quantum effect (Ryder, 1964; Bongaarts and Feeny, 1998).

TFR reflects fertility experience of women of childbearing age or women biologically capable of producing a birth. However, only those women can deliver birth who are sexually active. In India, sexual activity outside the institution of marriage is not socially accepted and, therefore, virtually all births in the country are confined within the institution of marriage. The latest (2019-2021) round of NFHS informs that around 1.268 million births that were reported by women of childbearing age in India during one year before the survey, only 684 births were reported by those women who were not married at the time of the survey. This means that TFR is influenced by both fertility of married of women of childbearing age and proportion of married women in different ages of the childbearing period. There may be a possibility that TFR may decrease despite the increase in the fertility of married women because of the decrease in the proportion of married women and vice versa.

Estimates of total marital fertility rate (TMFR) in India are available from the official sample registration system only. These estimates suggest that TMFR, in India, decreased from about 5.5 births per married woman of childbearing age during 1985-1987 to around 4.2 births per married women of childbearing age during 2012-2014 but then increased to around 5.1 births per married women of childbearing age during 2018-2020. During 1985-2014, the decrease in TFR in India has been associated with the decrease in TMFR but TFR decreased during 2012-2020 despite an increase in TMFR.

Both TFR and TMFR are composite measures of the fertility experience of women or married women of childbearing age. TFR is the simple arithmetic mean of age-specific fertility rates multiplied by 35, the duration of the childbearing period. Similarly, TMFR is 35 times the simple arithmetic mean of age-specific marital fertility rates. In this conceptualisation, TFR and TMFR are based on the simple arithmetic mean as the aggregation function to aggregate the fertility experience respectively of women and married women. The use of simple arithmetic mean as the aggregation function for the construction of composite measures has, however, been widely discussed and debated, especially, in the context of the human development index as it has certain inherent weaknesses (Desai, 1991, Kovacevic, 2010; Klugman et al, 2011). A major weakness of the simple arithmetic mean as the aggregation function to aggregate fertility experience of women/married women is that it embodies perfect substitutability of age-specific fertility/marital fertility rates. Another problem is that since fertility/marital fertility varies by age, the contribution of fertility/marital fertility rate in different ages to the simple arithmetic mean is different for different – the higher the fertility/marital fertility in an age, the higher its contribution to the simple arithmetic mean of age-specific fertility/ marital fertility rates. This implies that the contribution of the change in age-specific fertility/ marital fertility rates to the change in their simple arithmetic mean varies by age – the higher the fertility/marital fertility, the larger the change and the larger the contribution to simple arithmetic mean. There may be a possibility that fertility/marital fertility increases in some ages but decreases in others so that there is no change in the simple arithmetic mean. There is also a possibility that the direction of the change in fertility/marital fertility in some ages is opposite to the of the change in the simple arithmetic mean. In such a situation, the change in simple arithmetic mean reflects neither the increase in fertility/marital fertility in some ages or the decrease in fertility/marital fertility in other ages of the childbearing period. The age-specific fertility/marital fertility rates are ratios – number of births to number of women/married women in different ages of the childbearing period. Simple arithmetic mean is not appropriate to aggregate a set of ratios as the sum of the ratios is not equal to the ratio of the sum. Because of this flaw, both TFR and TMFR are interpreted in an hypothetical perspective only.

Alternatively, fertility experience of women/married women of childbearing age may be aggregated using the power or generalised mean with the power of the mean less than 1 (Bullen, 2000). When the power of the mean is equal to zero, the generalised mean is the geometric mean. The most important property of the geometric mean is that the geometric mean of a set of ratios is equal to the ratio of the geometric mean of the numerators of the ratios to the geometric mean of the denominators of the ratios. Geometric mean also addresses the problem of perfect substitutability associated with the simple arithmetic mean. The change in the geometric mean of age-specific fertility/ marital fertility rates gives equal weight to the change in fertility/marital fertility in different ages of the childbearing period. The geometric mean of the age-specific fertility/marital fertility rates is equal to the simple arithmetic mean of age-specific fertility/marital fertility rate only when fertility/marital fertility is the same in all ages of the childbearing period. Otherwise, the geometric mean is always less than the simple arithmetic mean of age-specific fertility/marital fertility rates. The ratio of the simple arithmetic mean to the geometric mean of the age-specific fertility/marital fertility rates, therefore, reflects the variation in fertility/marital fertility by age – the higher the ratio the larger the variation in fertility/marital fertility by age across the childbearing period. The increase in this ratio, therefore, implies an increase in the inequality of fertility/marital fertility across ages and vice versa. The concentration of fertility/marital fertility, in the younger ages of the childbearing period along with the decrease in fertility has implications for population stabilisation. From the perspective of population stabilisation, it is imperative that, with the transition, fertility/marital fertility should be more evenly distributed across the childbearing period, This means that the ratio of the simple arithmetic mean to the geometric mean of age-specific fertility/marital fertility rates should decrease with fertility transition.

In this paper, we show that fertility transition in India during 1985-2020 has been different when reflected through the trend in the simple arithmetic mean of age-specific fertility/marital fertility rates as compared to when it is reflected through the trend in the geometric mean of age-specific fertility/marital fertility rates. The paper shows that fertility transition is contingent upon how the age-specific fertility/marital fertility rates are aggregated. Our analysis shows that it is not appropriate to analyse fertility transition in terms of the trend in the simple arithmetic mean of age-specific fertility/marital fertility rates or, equivalently, in terms of TFR/TMFR. Rather, fertility transition should be analysed in terms of the trend in the geometric mean of age-specific fertility/marital fertility rates.

The rest of the paper is organised into seven sections. The next section outlines the methods used for analysing fertility transition. Our main assumption is that the trend in simple arithmetic mean/ geometric mean of age-specific fertility/marital fertility rates during 1985-2020 has changed at least once so that the entire trend can be divided into more than one time-segment and the trend in different time-segments may be different. The third section of the paper describes the data source. We have used the data from the official sample registration system of the country which is the only source that gives annual estimates of age-specific fertility/marital fertility rates. The fourth section presents results of the trend analysis while the fifth section factors the change in the simple arithmetic mean/geometric mean of age-specific fertility rates into the change in age-specific marital fertility rates and proportion of married women in different ages of the childbearing period. The sixth section presents the trend in the ratio of the simple arithmetic mean to the geometric mean of the age-specific fertility rates and the mean age of childbearing to analyse the change in age pattern of fertility. The last section summarises the findings of the analysis and their policy and programme implications in the context of population stabilisation.

## Methods

Let *f*_*i*_ denotes the fertility rate in age i of the childbearing period. Then, *f*_*i*_ can be summarised in terms of both simple arithmetic mean, *f*_*a*_, and geometric mean, *f*_*g*_, of *f*_*i*_. Or

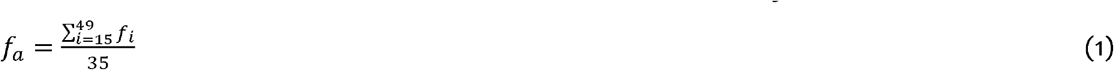

and

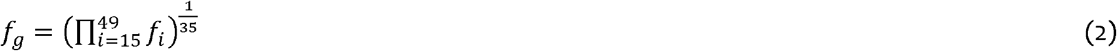

Here 35 is the length of the childbearing period. Similarly, if *g*_*i*_ denotes the fertility of married women in age i, then *g*_*i*_ can be summarised in terms of simple arithmetic mean, *g*_*a*_, and geometric mean, *g*_*g*_. of *g*_*i*_.

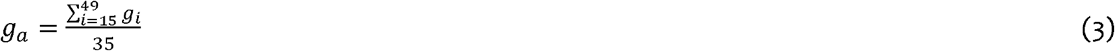

and

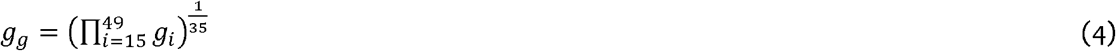

Notice than mi=*f*_*i*_/*g*_*i*_ is, by definition, the proportion of married women in age i. We analyse fertility transition in terms of the trend in *f*_*a*_ and *f*_*g*_ assuming that the trend is not linear but might have changed at least once so that the entire trend period can be divided into more than one time-segments of varying length with different trend in different time-segments. The first step in the trend analysis, therefore, is to identify the time(s) or the year(s) when the trend has changed or the joinpoint(s). The annual per cent change (APC) in different time-segment may then be calculated assuming a linear trend within the time-segment. The APC in different time-segments has then be aggregated into the average annual per cent change (AAPC) in the entire trend period as the weighted average of APC with weights proportional to the length of time-segments (Clegg et al, 2009). This approach best summarises the trend that varies over time (Marriot, 2010).

Several methods have been proposed to statistically determine the number of times the trend has changed. These include permutation test method (Kim et al, 2000); Bayesian Information Criterion (BIC) method (Kim et al, 2009); BIC3 method (Kim and Kim, 2016); modified BIC method (Zhang and Siegmund, 2007); weighted BIC method; and data dependent selection method (Kim et al, 2022). The permutation method is the gold standard. It uses the sequence of permutation tests to ensure that the approximate probability of the overall Type I error is less than the specified significance level. Assuming that the default value of the minimum number of joinpoint(s) is 0, “the overall Type I error” is the probability of incorrectly concluding that the underlying model has at least one joinpoint when, in fact, the true underlying model has no joinpoint. In the present analysis, we have used the data dependent selection method to identify joinpoint(s). This method internally determines the model selection, BIC or BIC3, based on the characteristics of the data. The basic idea is to use BIC if change sizes are relatively small and BIC3 otherwise.

Actual calculations have been caried out using the open-source software Joinpoint Regression Program (National Cancer Institute, 2024). The software requires specification of minimum (0) and maximum number of joinpoints (>0) in advance. We have specified the minimum number of joinpoints as 0 and the maximum number of joinpoints has been set to 5. The programme starts with the minimum number of joinpoints and tests whether more joinpoints are statistically significant and must be added to the model (up to the pre-specified maximum number of joinpoints). The tests of significance are based on a Monte Carlo Permutation method (Kim et al, 2000). The grid search method has been used to identify joinpoints (Lerman, 1980). This method allows a joinpoint to occur exactly at time t. A grid is created for all possible positions of the joinpoint(s) or of the combination of joinpoint(s) and then the model is fitted for each possible position of the joinpoint(s), Finally, that position of joinpoint(s) is selected which minimises the sum of squared errors (SSE).

The change in *f*_*a*_ between time *t*_1_ and *t*_2_ (*t*_2_>*t*_1_) can be factored into the change in *g_i_* and the change in m_i_. We can write

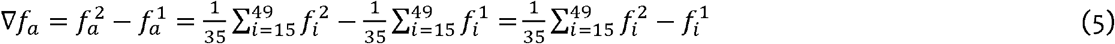

Now

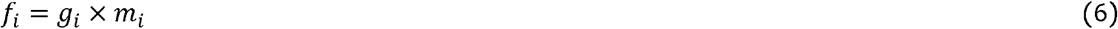

so that

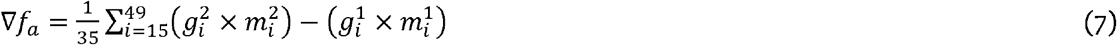

Now

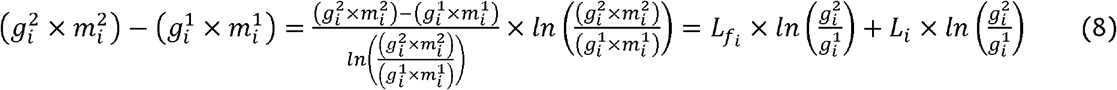

Where

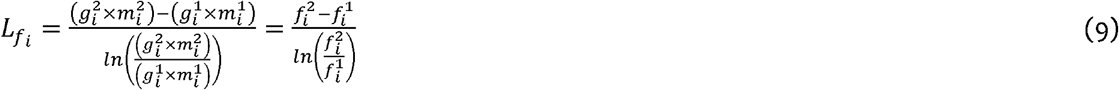

is the logarithmic mean of *f*_*i*_ at times *t*_1_ and *t*_2_ and *t*_2_>*t*_1_ (Bhatia, 2008; Carlson, 1966). In other words,

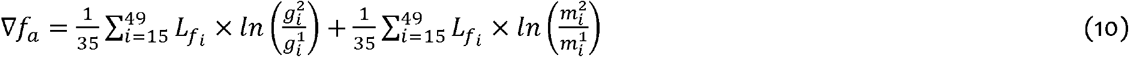

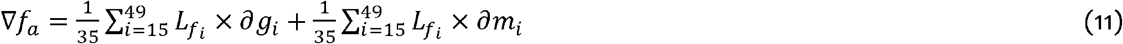

Where

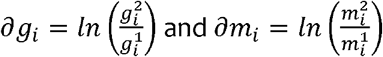

Similarly, the change in *f*_*g*_, between time *t*_1_ and *t*_2_ (*t*_2_>*t*_1_) can be written as

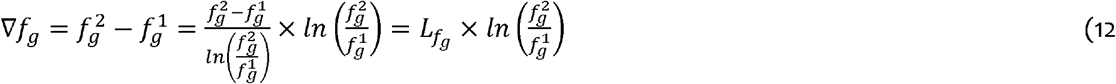

Where

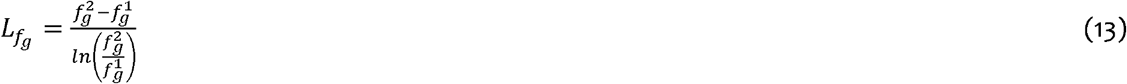

is the logarithmic mean of *f*_*g*_ at times *t*_1_ and *t*_2_ (*t*_2_>*t*_1_). Now

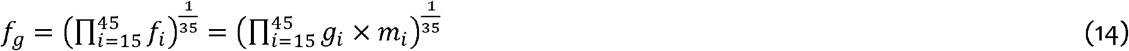

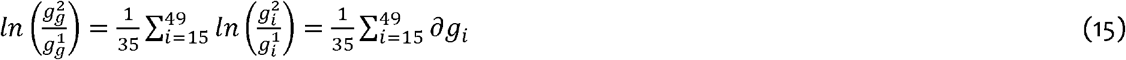

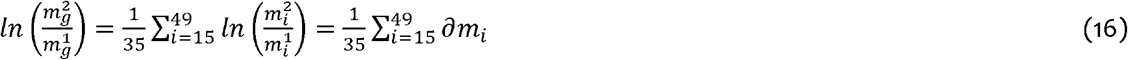

so that

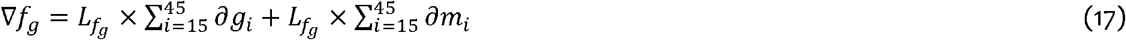

The decomposition of the change in *f*_*g*_ given by equation (17) is different from the decomposition of the change in fa given by equation (10) in terms of the multiplying factor of the change in the marital fertility rate and the change in the proportion of married women in different ages of the childbearing period. In the decomposition of the change in *f*_*g*_ given by equation (20), this multiplication factor is the same for all ages of the childbearing period. In the decomposition of the change in *f*_*a*_ given by equation (11), this multiplying factor varies by age. The change in *f*_*a*_ gives more importance to the change in marital fertility and the change in the proportion of married women in those ages in which fertility is high as compared to those ages in which fertility is low. This means that the analysis of the change in fertility in terms of the trend in the simple arithmetic mean of age-specific fertility rates, *f*_*a*_, is biased towards the change in those ages in which fertility is high relative to those ages in which fertility is low. This is not the case with the analysis of the change in fertility in terms of the trend in the geometric mean of age-specific fertility rates, *f*_*g*_, which gives equal importance to the change in fertility in different ages of the childbearing period. Since both marital fertility and the proportion of married women vary by age, it is logical to argue that the analysis of fertility transition should give equal weight to the change in fertility and the change in the proportion of married women in different ages of the childbearing period. This is possible only when the fertility experience of women of childbearing age is summarised or the age-specific fertility rates are aggregated using the geometric mean of age-specific fertility rates rather than the simple arithmetic mean of the age-specific fertility rates. The trend in the simple arithmetic mean of age-specific fertility rates and hence in the total fertility rate is biased towards the trend in fertility in those ages of the childbearing period in which fertility is high compared to ages in which fertility is low. It is imperative in the analysis of fertility transition that importance is given to fertility change in different ages of the childbearing period irrespective of the level of fertility. For this very reason, the trend in the simple arithmetic mean age-specific fertility rates or in total fertility rate may depict a misleading picture of fertility transition. It is more appropriate to use the geometric mean of age-specific fertility rates to analyse fertility transition.

### Data

The analysis is based on the annual estimates of age-specific fertility rates and age-specific marital fertility rates available from the official sample registration system of India. The sample registration system was launched by the Government of India on a pilot basis in 1964-1965 and was expanded to cover the entire country in 1969-1970. It is a large-scale survey of a sample of households in the country. The revision the sampling frame is carried out after every decennial population census in the country. The first revision of the sampling frame was carried out in 1977-1978 while the last revision was carried out in 2014. In 1969-1970, the system covered 3722 sampling units throughout the country. This number has increased to 8853 in 2014 (Government of India, 2022b). The households for the survey under the system are selected through a uni-stage, stratified simple random sampling without replacement approach with some variations in specific cases. In 2020, the system covered more than 8.3 million population throughout the country. The system collects information about the vital events in the household through a dual record system - continuous enumeration of births and deaths in the sampled villages/urban blocks by a resident part-time enumerator, and an independent six-monthly retrospective survey covering all households in the village/urban block by a full-time supervisor. The information about vital events obtained from the two sources are matched and the unmatched and partially matched vital events are re-verified in the field to get an unduplicated count of the correct number of vital events in the village/urban block. In addition, a base-line survey is also carried out prior to the start of continuous enumeration in each sampled village/block to prepare a notional map of the area to be surveyed, and for house numbering and house listing (Government of India, 2022a).

The sample registration system is the only system in India which provides annual estimates of selected indicators of fertility and mortality for the country and for the constituent states and Union Territories of the country. The system is, however, not designed to provide district level estimates of the indicators of fertility and mortality. The registration of births and deaths in India are mandatory under the Registration of Births and Deaths Act of 1969 which has been amended recently in 2023 (Government of India, 1969; 2023). However, the completeness of birth and death registration in the country remains far from satisfactory to provide statically reliable estimates of indicators of fertility and mortality. Estimates of the indicators of fertility in India are also available from the National Family Health Survey (NFHS) Programme launched by the Government of India in 1992-1993. The latest round of NFHS was carried out in 2019-2021 (Government of India, 2022b). The NFHS, however, is not carried out annually d, therefore, does not provide estimates of selected demographic indicators on an annual basis.

Annual estimates of the birth rate, age-specific fertility rates and the total fertility rate (TFR) in India are available from the sample registration system for the period 1970-2020. However, estimates of age-specific marital fertility rate and total marital fertility rate (TMFR) are available for the period 1985-2020 only. The present analysis has, therefore, been confined to the analysis of the change in fertility in the country during the period 1985-2020 only. Estimates of demographic indicators available from the sample registration system are also known to be associated with random errors of unknown origin. It is, therefore, customary to use three-year moving average in place of the annual estimates of demographic indicators available from the system for the analysis of the trend in indicators. We have also followed the same convention in the present analysis. Three-years moving average has been used to iron out the random errors of unknown origin associated with the estimates of demographic indicators derived from the sample registration system. We have assumed that the three-year average of the demographic indictor is located at the mid-point of the three-year interval. For example, the average of the TFR for the years 1985, 1986 and 1987 available from the sample registration system is assumed to be the TFR for the year 1986. It may, however, be noticed that this average TFR for the three-year period may be different from the actual TFR derived directly from the data available from the system. The average of the TFR for the period 1985-1987 may not be the same as the TFR for the year 1987 estimated directly from data available from the system.

### Trend in *f*_*a*_ and *f*_*g*_

Figure 1 depicts the trend in the simple arithmetic mean (*f*_*a*_) and geometric mean (*f*_*g*_) of the age-specific fertility rates. The trend in the two aggregate measures of age-specific fertility rates is different. The trend in *f*_*a*_ or, equivalently in TFR, suggests that fertility in India has decreased consistently during the period 1985-2020 whereas the trend in *f*_*g*_ suggests that the decrease in fertility has stagnated after the period 2011-2013. Figure 1 shows that the trend in fertility is contingent upon the function used to aggregate the age-specific fertility rates. The arithmetic mean of age-specific fertility rates show an almost linear decrease in fertility whereas the geometric mean of age-specific fertility rates suggests that fertility in the country has stopped decreasing after 2011-2013 and has virtually remained unchanged during the period 2011-2020. Since, fertility rate is not the same in all ages of the childbearing period, but varies by age, the geometric mean is the more appropriate function to aggregate the age-specific fertility rates than the simple arithmetic mean of the age-specific fertility rates.

**Figure 1:**
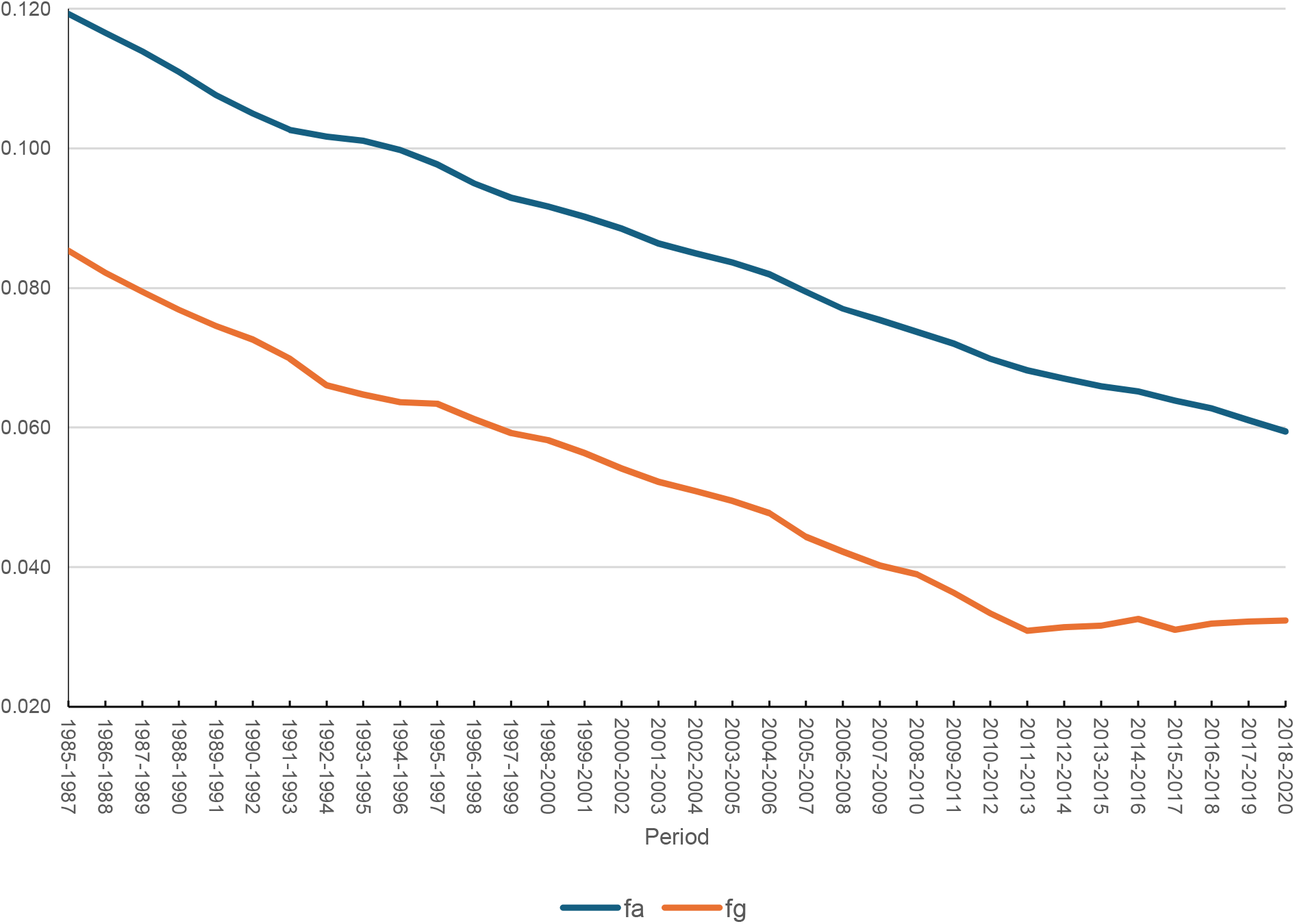
Trend in *f*_*a*_ and *f*_*g*_ in India, 1985-2020. Source: Author, based on the data from the sample registration system.

Figure 1 also indicates that the trend in fertility in the country has not been uniform during the period 1985-2020 irrespective of whether fertility is measured in terms of *f*_*a*_ or *f*_*g*_. identify the joinpoint(s) or the year(s) when the trend has changed, we have applied the joinpoint regression analysis of the trend in both *f*_*a*_ and *f*_*g*_ and the results are presented in table 1. The maximum number of joinpoints or the number of times the trend has changed was set to five the joinpoint regression analysis and the data driven selection method was used to identify the joinpoints or the time when the trend has changed. Table 1 reveals that the trend in both *f*_*a*_ and *f*_*g*_ changed at least five times during the trend period 1985-2020. The time when the trend in *f*_*a*_ change and the time when the trend in *f*_*g*_ changed was, however, different. Table 1 also reveals that the average annual per cent change (AAPC) during the period 1985-2020 in *f*_*a*_ and *f*_*g*_ has also been different. Table 1 confirms that the change in fertility reflected by the trend in the simple arithmetic mean of age-specific fertility rates, *f*_*a*_, is different from the change in fertility reflected by the trend in the geometric mean of age-specific fertility rates, *f*_*g*_. This difference in the trend in *f*_*a*_ and *f*_*g*_ is particularly marked during the period 2011-2020 when the annual per cent change (APC) in *f*_*a*_ was negative meaning a decrease in *f*_*a*_ but the APC in *f*_*g*_ was positive meaning an increasing *f*_*g*_ although the APC in *f*_*g*_ during 2011-2020 was not statistically insignificantly different from zero. Before the period 2011-2013, however, the trend in both *f*_*a*_ and *f*_*g*_ has been similar, although the times when the trend has changed in *f*_*a*_ are different from the times when the trend in *f*_*g*_ has changed and the annual per cent change (APC) in *f*_*a*_ and *f*_*g*_ during different time-segments has been different. Another observation of table 1 is that change in fertility reflected through the trend in *f*_*a*_ is slower than the change in fertility reflected through the trend in *f*_*g*_.

**Table 1:**
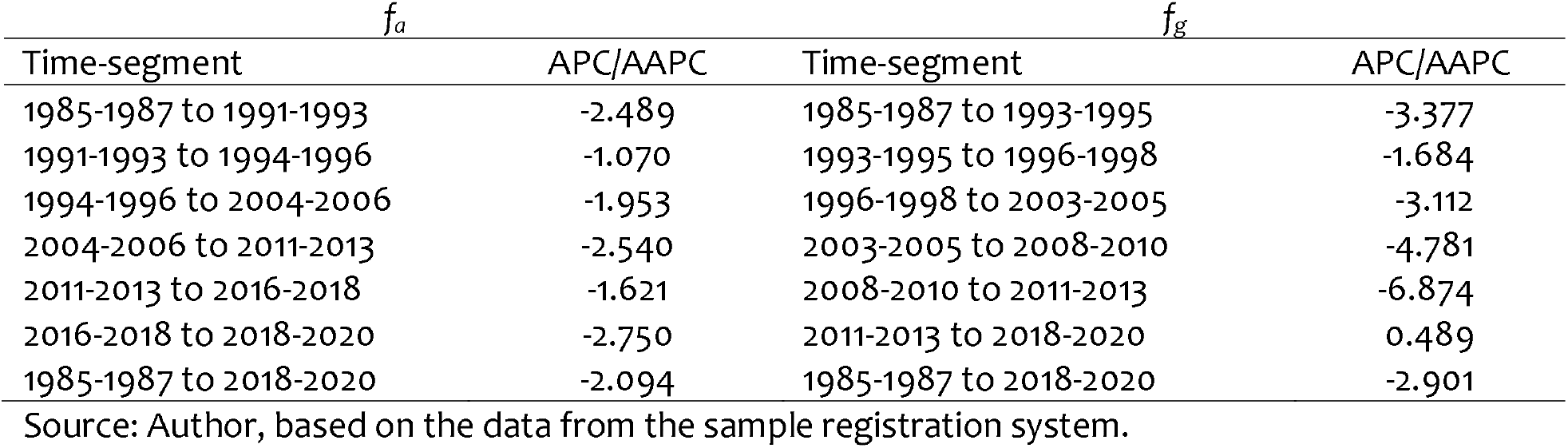
Annual per cent change (APC) in different time-segments and average annual per cent change (AAPC) during 1985-2020 in simple arithmetic mean (*f*_*a*_) and geometric mean (*f*_*g*_) of age-specific fertility rates in India.

| $f_a$ | | $f_g$ | |
| --- | --- | --- | --- |
| Time-segment | APC/AAPC | Time-segment | APC/AAPC |
| 1985-1987 to 1991-1993 | -2.489 | 1985-1987 to 1993-1995 | -3.377 |
| 1991-1993 to 1994-1996 | -1.070 | 1993-1995 to 1996-1998 | -1.684 |
| 1994-1996 to 2004-2006 | -1.953 | 1996-1998 to 2003-2005 | -3.112 |
| 2004-2006 to 2011-2013 | -2.540 | 2003-2005 to 2008-2010 | -4.781 |
| 2011-2013 to 2016-2018 | -1.621 | 2008-2010 to 2011-2013 | -6.874 |
| 2016-2018 to 2018-2020 | -2.750 | 2011-2013 to 2018-2020 | 0.489 |
| 1985-1987 to 2018-2020 | -2.094 | 1985-1987 to 2018-2020 | -2.901 |

Source: Author, based on the data from the sample registration system.

Table 1 also shows that the slowdown in the decrease in simple arithmetic mean of the age-specific fertility rates, *f*_*a*_, or, equivalently, in TFR during the time-segment 1991-1996 and again during the time-segment 2011-2018 was primarily responsible for the delay in achieving the replacement fertility in India. If the annual per cent change (APC) in *f*_*a*_ and hence in TFR observed during 1985-1993 would have been maintained during the post 1993 period, the TFR would have decreased to the replacement level by the year 2013 or about three years later than the target date set in the National Population Policy 2000. The decrease in *f*_*a*_, or in TFR, accelerated during the period 1994-2013 but the decrease in fertility decelerated again during the period 2011-2018 which also contributed for the delay in the achievement of replacement fertility in the country.

Table 1 confirms that the function used to aggregate age-specific fertility rates matters in analysing fertility change. When simple arithmetic mean is used, fertility in India appears to have decreased throughout the period 1985-2020. However, when geometric mean is used, decrease in fertility appears to have stalled during 2011-2020. The simple arithmetic mean embodies perfect substitutability. The rapid decrease in fertility in some ages appears to have offset increase in fertility in other ages so that *f*_*a*_, or TFR continued to decrease during 2011-2020 reflecting continued decrease in fertility in the country. In case of geometric mean, this substitution effect is minimal, although not fully eliminated so that the increase in fertility in some ages cannot be fully offset by the decrease in fertility in other ages. It is obvious that the difference in the change in fertility reflected by the trend in *f*_*a*_ and the trend in *f*_*g*_ is not because of the difference in the fertility experience of women of childbearing age but because of the limitation of the simple arithmetic mean to aggregate the age-specific fertility rates because the simple arithmetic mean embodies perfect substitutability.

The trend in marital fertility has also been found to be sensitive to selection of the aggregation function (Table 2). The average annual per cent change (AAPC) in the simple arithmetic mean of age-specific marital fertility rates, *g*_*a*_, was -0.280 per cent during 1985-2020 but -1.478 per cent in the geometric mean of age-specific marital fertility rates. The sensitiveness of the trend in marital fertility to the aggregation function used to aggregate the age-specific marital fertility rates is obvious. The trend in the simple arithmetic mean of age-specific marital fertility rates, *g*_*a*_, and the geometric mean of age-specific marital fertility rates, *g*_*g*_, has also been different as may be seen from the times when the trend has changed and the annual per cent change (APC) in *g*_*a*_ and *g*_*g*_ in different time-segments of the period 1985-2020.

**Table 2:**
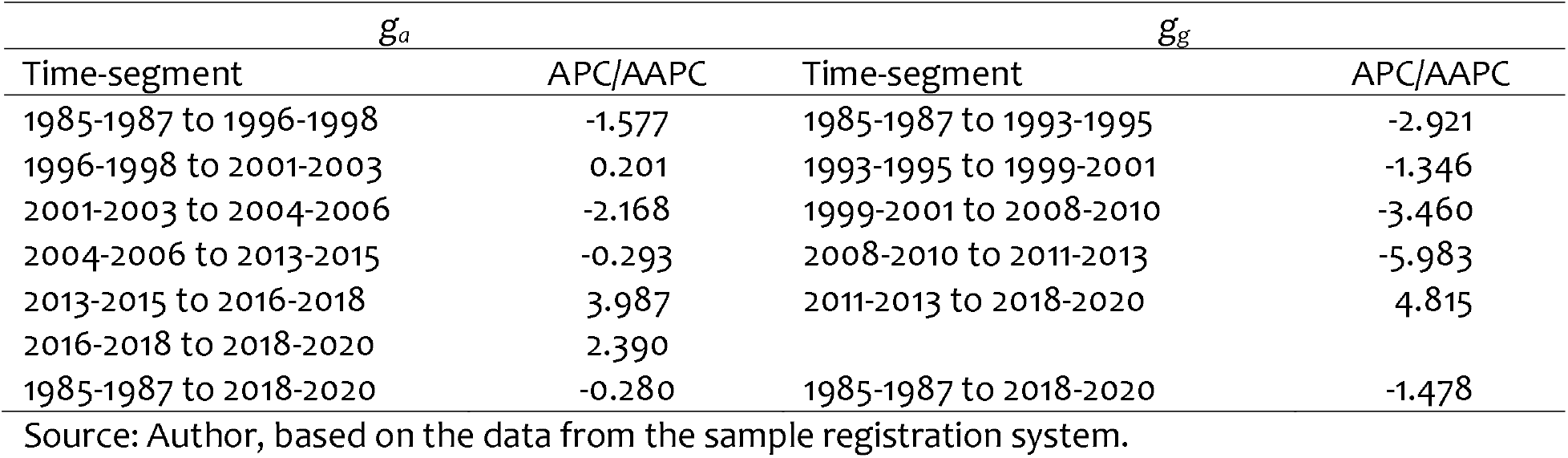
Annual per cent change (APC) in different time-segments and average annual per cent change (AAPC) during 1985-2020 in simple arithmetic mean, *g*_*a*_, and geometric mean, *g*_*g*_, of age-specific marital fertility rates in India, 1985-2020.

### Trend in Age-specific Fertility/Marital Fertility Rates

The sensitiveness of the change in fertility to the aggregation function used for aggregate age-specific fertility rates – simple arithmetic mean or geometric mean - suggests that any analysis of the change in fertility based on the trend in the aggregate measures of age-specific fertility rates may be fraught with problems and may even be misleading. Otherwise also, the trend in aggregate measures of age-specific fertility rates presents only a rounded assessment of the change in fertility and masks the change in age-specific fertility rates. There may be a situation where the direction of the change in age-specific fertility rates is not the same and the direction of the change in fertility rate in some ages is opposite to the direction of the change in the aggregate measure of fertility. It is, therefore, imperative for a fuller understanding of the change in fertility that the analysis of the trend in the aggregate measure of fertility is accompanied with the analysis of trend in age-specific fertility rates. The analysis of the trend in the age-specific fertility rates is necessary to avoid the pitfalls in interpreting the change in fertility, especially when the simple arithmetic mean is used as the aggregation function.

Figure 3 depicts the trend in fertility rates in the conventional five years age-groups in India during the period 1985-2020. The most revealing observation of the figure 3 is that the trend in different age-specific fertility rates in the country has not been the same, especially, after the period 2012-2014. Fertility has decreased in some age-groups during the period 2012-2020 but has increased in other age-groups. There has been a marked decrease in fertility of women aged 20-24 years but a marked increase in fertility of women aged 15-29 years after 2012-2014. D has been different prior to 2012-2014 as fertility decreased in all age-groups, although the pace of decrease in fertility has varied by age. The trend in age-specific fertility rates has also not been linear during the period 1985-2020 but has changed at least once so that the pace of change has been different in different time-segments. The joinpoint regression analysis has been used to identify the time when the trend has changes and to calculate annual per cent change (APC) in different time-segments.

**Figure 2:**
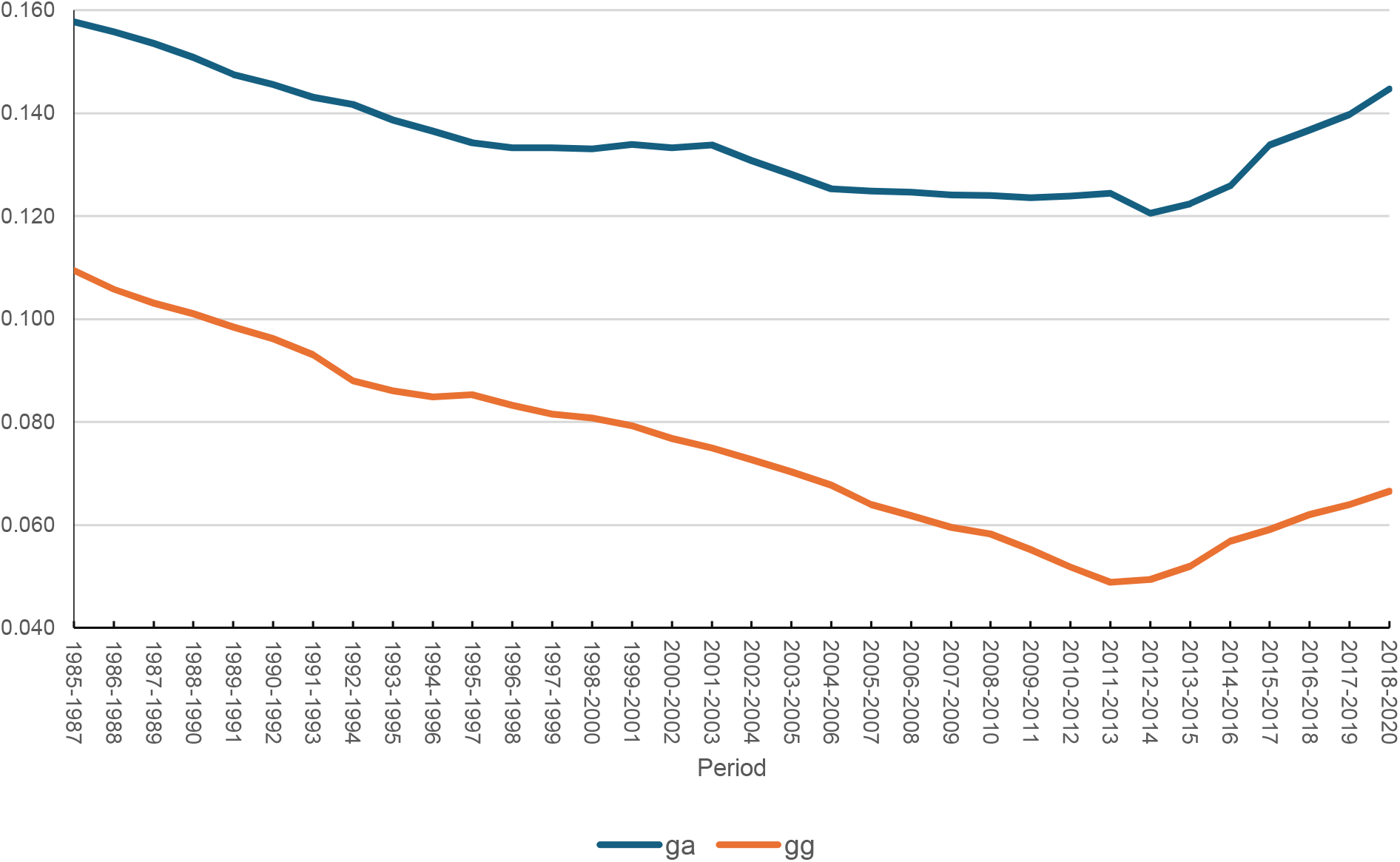
Trend in simple arithmetic mean (*g*_a_) and geometric mean (*g*_*g*_) of age-specific marital fertility rates in India, 1985-2020. Source: Author, based on the data from the sample registration system.

**Figure 3:**
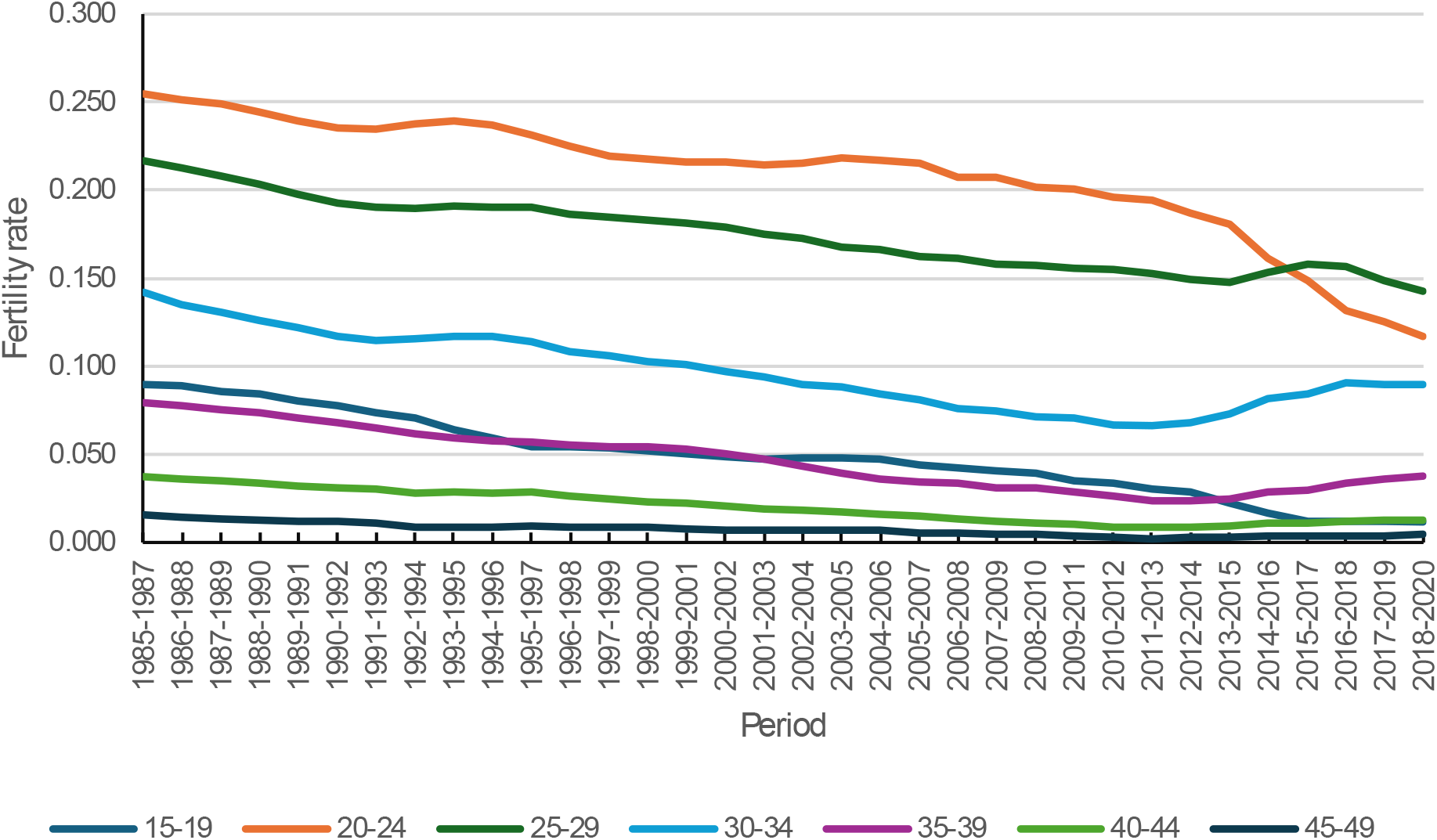
Trend in age-specific fertility rates in India, 1985-2020. Source: Author, based on the estimates available from the sample registration system.

Results of the joinpoint regression analysis of the trend in age-specific fertility rates and age-specific marital fertility rates are presented in table 3. The change in fertility has been the fastest in the age-group 15-19 years. Fertility decreased at an average annual per cent decrease of more than 6 per cent during the period 1985-2020 in this age group. This age-group is the only age-group in which fertility decreased in all time-segments of the period 1985-2018 identified through the joinpoint regression analysis. In all other age-groups, fertility increased at least in one time-segment of the period 1985-2020. The table also shows that fertility increased quite rapidly in the ages 30-49 years after 2011-2013 with the only exception of decrease in fertility in the age group 30-34 years during 2016-2020.

**Table 3:**
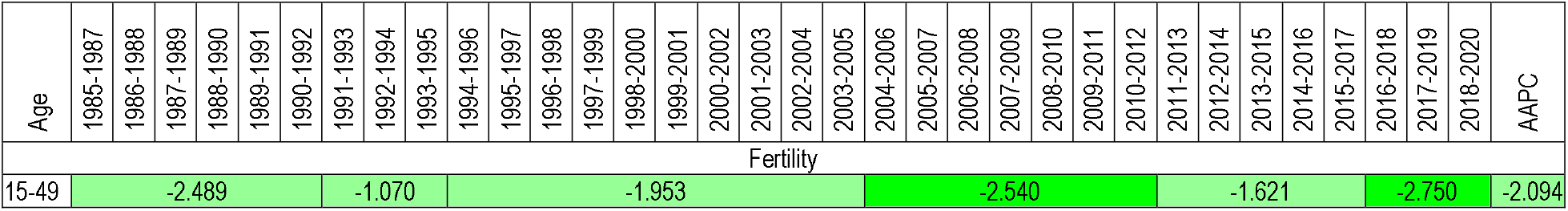
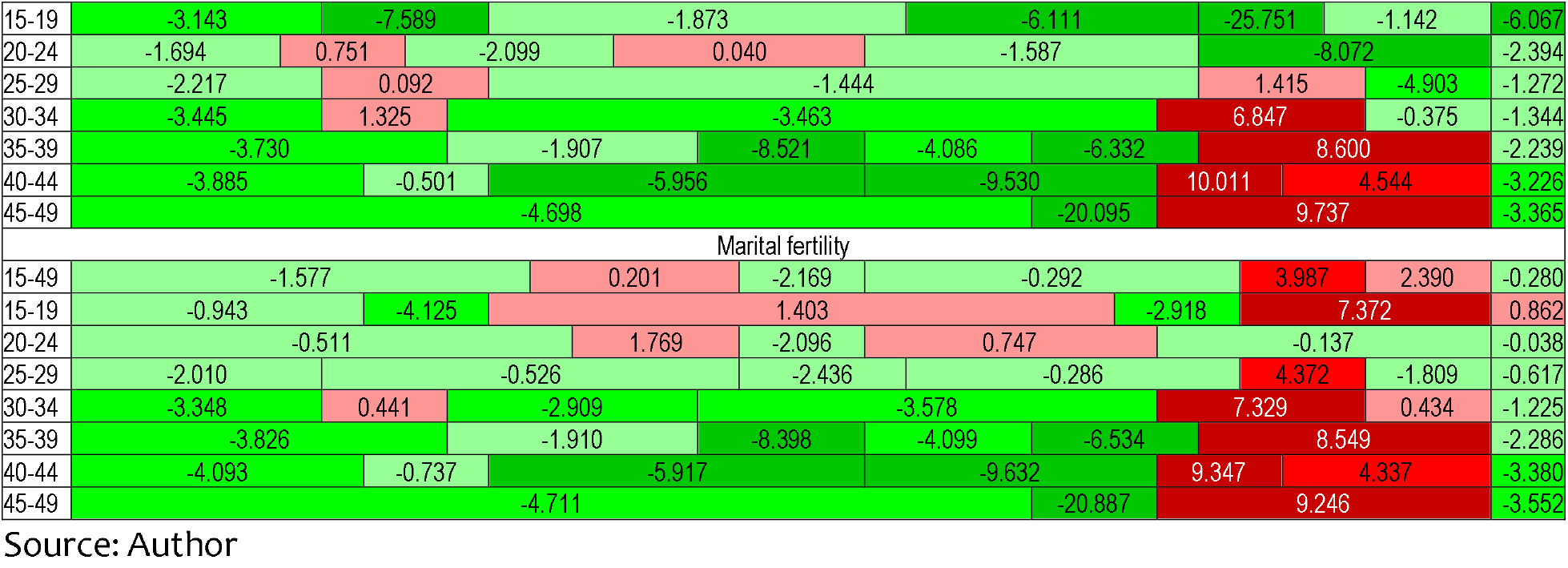
Average annual per cent change (AAPC) during 1985-2020 and annual per cent change (APC) in different time segments in age-specific fertility rates and age-specific marital fertility rates in India.

On the other hand, there is no age-group in which fertility of married women decreased in all time-segments of the period 1985-2020 identified through the joinpoint regression analysis. Marital fertility increased very rapidly during the time-segment 2013-2020 in the age group 15-19 years so that the marital fertility in this age group increased, instead decreased during the entire trend period 1985-2020. The average annual per cent change (decrease) in fertility has been faster than the average annual change in marital fertility in the age-group 15-39 years but, in the age group 40-49 years, the average annual per cent change in marital fertility has been faster than that in fertility.

Table 3 also confirms that the change in fertility in India has been different during the period 1985-2012 and during the period 2011-2020. During the period 1985-2012, fertility decreased in all ages of the childbearing period with only a few exceptions, although with varying pace of decrease. However, during the period 2011-2020, fertility, in general, increased in the age-group 25-49 years with some exceptions and the increase in fertility has been quite rapid. During the post 2011-2013 period, fertility transition in the country has virtually been confined to the younger ages of the childbearing period only. In the older ages of the childbearing period, there has apparently been a reversal in fertility transition in the post 2011-2013 period. In case of marital fertility, on the other hand, the reversal in fertility transition in almost all ages of the childbearing period is very much evident from table 3. Marital fertility decreased in the age group 20-24 years only during the post 2011-2013 period. The marital fertility in the age group 15-19 years increased during the time-segment 1995-2011 and again during the time-segment 2013-2020 so that marital fertility in this age-group increased, instead decreased during the entire trend period 1985-2020.

### Decomposition of the Change in Fertility

The change in both *f*_*a*_ or *f*_*g*_ is due to the change in marital fertility rate and the change in the proportion of married women in different ages. Table 4 gives the change in the age-specific marital fertility rates (*g*_*i*_) and the change in the age-specific proportion of married women (mi) during 1985-2020. The marital fertility increased, instead decreased, in the age-group 15-24 years but decreased in other age-groups during the period 1985-2020 and the decrease was the most rapid in the age group 40-49 years. The change in marital fertility in different age-groups was, however, different in different time-segments. It was during 1985-1993 only when marital fertility decreased in all ages of the childbearing period. By contrast, marital fertility increased in all ages of the childbearing period, except 20-24 years, during 2011-2018. Similarly, marital fertility also increased in all ages of the childbearing period, except 25-29 years, during 2016-2020. In both these time-segments, increase in marital fertility was the most rapid in the age group 45-49 years.

**Table 4:** Decomposition of the change in the simple arithmetic mean of age-specific fertility rates, *f*_*a*_, in India, 1985-2020.

| Time-segment | Age group |  |  |  |  |  |  |
| --- | --- | --- | --- | --- | --- | --- | --- |
|  | 15-19 | 20-24 | 25-29 | 30-34 | 35-39 | 40-44 | 45-49 |
| Change in marital fertility ($\partial g_i$) | | | | | | | |
| 1985-1987 to 1991-1993 | -0.0448 | -0.0260 | -0.1132 | -0.2076 | -0.2111 | -0.2414 | -0.2976 |
| 1991-1993 to 1994-1996 | -0.1120 | -0.0153 | -0.0165 | 0.0117 | -0.1199 | -0.0678 | -0.3241 |
| 1994-1996 to 2004-2006 | 0.0811 | -0.0108 | -0.1109 | -0.3200 | -0.4555 | -0.5572 | -0.2060 |
| 2004-2006 to 2011-2013 | 0.1010 | 0.0678 | -0.0327 | -0.2335 | -0.4123 | -0.6987 | -1.0818 |
| 2011-2013 to 2016-2018 | 0.1412 | -0.0498 | 0.1067 | 0.3233 | 0.3159 | 0.3413 | 0.5008 |
| 2016-2018 to 2018-2020 | 0.1208 | 0.0454 | -0.0226 | 0.0165 | 0.1183 | 0.0790 | 0.1345 |
| 1985-1987 to 2018-2020 | 0.2873 | 0.0112 | -0.1892 | -0.4097 | -0.7646 | -1.1449 | -1.2743 |
| Change in proportion of married women ($\partial m_i$) | | | | | | | |
| 1985-1987 to 1991-1993 | -0.1521 | -0.0554 | -0.0157 | -0.0030 | 0.0037 | 0.0112 | -0.0444 |
| 1991-1993 to 1994-1996 | -0.0969 | 0.0273 | 0.0176 | 0.0065 | 0.0035 | 0.0071 | 0.0244 |
| 1994-1996 to 2004-2006 | -0.3055 | -0.0807 | -0.0230 | -0.0082 | -0.0068 | -0.0003 | -0.0081 |
| 2004-2006 to 2011-2013 | -0.5587 | -0.1765 | -0.0549 | -0.0086 | 0.0045 | 0.0076 | 0.0270 |
| 2011-2013 to 2016-2018 | -1.0636 | -0.3383 | -0.0813 | -0.0115 | 0.0055 | 0.0208 | 0.0214 |
| 2016-2018 to 2018-2020 | -0.1722 | -0.1634 | -0.0704 | -0.0180 | 0.0007 | 0.0034 | 0.0148 |
| 1985-1987 to 2018-2020 | -2.3490 | -0.7869 | -0.2277 | -0.0429 | 0.0110 | 0.0498 | 0.0352 |

Source: Author

On the other hand, the proportion of married women decreased during 1985-2020 in ages below 35 years but increased in ages 35 years and above. The decrease in the proportion of married women was very rapid in the age-group 15-19 years and decreased in all time-segments. This has, however, not been the case in other age-groups. The time-segment 1994-2006 is the only time-segment in which the proportion of married women decreased in all age-groups of the childbearing period whereas the time-segment 1991-1996 is the only time-segment in which the proportion of married women increased in all age-groups except the age group 15-19 years.

The contribution of the change in fertility and the change in the proportion of married women to the change in the arithmetic mean of age-specific fertility rates (*f*_*a*_) and the geometric mean of the age specific fertility rates (*f*_*g*_) has been different. The change in marital fertility during 1985-2020 accounted for a change of 35 per cent in *f*_*a*_ which means that the change in the proportion of married women accounted for a change of almost 65 per cent of the change in fa (Table 5). On the other hand, the change in marital fertility during 1985-2020 accounted for more than 51 per cent in *f*_*g*_ so that the change in the proportion of married women accounted for only about 49 per cent of the change in *f*_*g*_ (Table 6). Summarising the fertility experience of women of childbearing age through the simple arithmetic mean of the age-specific fertility rates leads to the conclusion that most of the decrease in fertility has been the result of the decrease in the proportion of married women while the decrease in the marital fertility has played only a secondary role. However, when the fertility experience of women of childbearing age is summarised through the geometric mean of age-specific fertility rates, both the decrease in the marital fertility and the decrease in the proportion of married women have contributed almost equally to the decrease in fertility. This shows that the contribution of the change in fertility to the change in fertility is also influenced by the selection of the aggregation function for summarising the fertility experience of women of childbearing age.

**Table 5:** Decomposition of the change in the simple arithmetic mean of age-specific fertility rates, *f*_*a*_, in India, 1985-2020.

| Time-segment | Age group | | | | | | | Change in $f_a$ |
| --- | --- | --- | --- | --- | --- | --- | --- | --- |
|  | 15-19 | 20-24 | 25-29 | 30-34 | 35-39 | 40-44 | 45-49 |  |
| 1985-1987 to 1991-1993 | -0.0160 | -0.0199 | -0.0262 | -0.0269 | -0.0149 | -0.0078 | -0.0046 | -0.1162 |
| 1991-1993 to 1994-1996 | -0.0138 | 0.0028 | 0.0002 | 0.0021 | -0.0071 | -0.0018 | -0.0029 | -0.0205 |
| 1994-1996 to 2004-2006 | -0.0120 | -0.0208 | -0.0239 | -0.0327 | -0.0213 | -0.0121 | -0.0016 | -0.1243 |
| 2004-2006 to 2011-2013 | -0.0175 | -0.0223 | -0.0140 | -0.0181 | -0.0121 | -0.0081 | -0.0044 | -0.0964 |
| 2011-2013 to 2016-2018 | -0.0181 | -0.0625 | 0.0039 | 0.0242 | 0.0091 | 0.0035 | 0.0016 | -0.0383 |
| 2016-2018 to 2018-2020 | -0.0006 | -0.0147 | -0.0139 | -0.0001 | 0.0042 | 0.0010 | 0.0006 | -0.0235 |
| 1985-1987 to 2018-2020 | -0.0780 | -0.1373 | -0.0738 | -0.0516 | -0.0421 | -0.0251 | -0.0112 | -0.4190 |
| Contribution of the change in marital fertility |  |  |  |  |  |  |  |  |
| 1985-1987 to 1991-1993 | -0.0036 | -0.0064 | -0.0230 | -0.0265 | -0.0152 | -0.0081 | -0.0040 | -0.0868 |
| 1991-1993 to 1994-1996 | -0.0074 | -0.0036 | -0.0031 | 0.0014 | -0.0073 | -0.0020 | -0.0031 | -0.0253 |
| 1994-1996 to 2004-2006 | 0.0043 | -0.0025 | -0.0198 | -0.0319 | -0.0210 | -0.0121 | -0.0015 | -0.0844 |
| 2004-2006 to 2011-2013 | 0.0039 | 0.0139 | -0.0052 | -0.0175 | -0.0123 | -0.0082 | -0.0045 | -0.0298 |
| 2011-2013 to 2016-2018 | 0.0028 | -0.0080 | 0.0165 | 0.0251 | 0.0090 | 0.0033 | 0.0015 | 0.0501 |
| 2016-2018 to 2018-2020 | 0.0014 | 0.0056 | -0.0034 | 0.0015 | 0.0042 | 0.0010 | 0.0006 | 0.0109 |
| 1985-1987 to 2018-2020 | 0.0109 | 0.0020 | -0.0335 | -0.0467 | -0.0427 | -0.0263 | -0.0115 | -0.1478 |
| Contribution of the change in the proportion of married women |  |  |  |  |  |  |  |  |
| 1985-1987 to 1991-1993 | -0.0123 | -0.0135 | -0.0032 | -0.0004 | 0.0003 | 0.0004 | -0.0006 | -0.0294 |
| 1991-1993 to 1994-1996 | -0.0064 | 0.0065 | 0.0033 | 0.0007 | 0.0002 | 0.0002 | 0.0002 | 0.0048 |
| 1994-1996 to 2004-2006 | -0.0163 | -0.0183 | -0.0041 | -0.0008 | -0.0003 | 0.0000 | -0.0001 | -0.0399 |
| 2004-2006 to 2011-2013 | -0.0213 | -0.0362 | -0.0088 | -0.0006 | 0.0001 | 0.0001 | 0.0001 | -0.0666 |
| 2011-2013 to 2016-2018 | -0.0209 | -0.0545 | -0.0126 | -0.0009 | 0.0002 | 0.0002 | 0.0001 | -0.0884 |
| 2016-2018 to 2018-2020 | -0.0020 | -0.0203 | -0.0105 | -0.0016 | 0.0000 | 0.0000 | 0.0001 | -0.0343 |
| 1985-1987 to 2018-2020 | -0.0888 | -0.1393 | -0.0403 | -0.0049 | 0.0006 | 0.0011 | 0.0003 | -0.2712 |

Source: Author

**Table 6:** Decomposition of the change in the geometric mean of age-specific fertility rates, *f*_*g*_, in India, 1985-2020.

| Time-segment | Age group | | | | | | | Change<br>in $f_g$ |
| --- | --- | --- | --- | --- | --- | --- | --- | --- |
|  | 15-19 | 20-24 | 25-29 | 30-34 | 35-39 | 40-44 | 45-49 |  |
| Change in fertility |  |  |  |  |  |  |  |  |
| 1985-1987 to 1991-1993 | -0.0022 | -0.0009 | -0.0014 | -0.0023 | -0.0023 | -0.0025 | -0.0038 | -0.0154 |
| 1991-1993 to 1994-1996 | -0.0020 | 0.0001 | 0.0000 | 0.0002 | -0.0011 | -0.0006 | -0.0029 | -0.0062 |
| 1994-1996 to 2004-2006 | -0.0018 | -0.0007 | -0.0011 | -0.0026 | -0.0037 | -0.0044 | -0.0017 | -0.0159 |
| 2004-2006 to 2011-2013 | -0.0025 | -0.0006 | -0.0005 | -0.0013 | -0.0023 | -0.0038 | -0.0058 | -0.0169 |
| 2011-2013 to 2016-2018 | -0.0041 | -0.0017 | 0.0001 | 0.0014 | 0.0014 | 0.0016 | 0.0023 | 0.0010 |
| 2016-2018 to 2018-2020 | -0.0002 | -0.0005 | -0.0004 | 0.0000 | 0.0005 | 0.0004 | 0.0007 | 0.0004 |
| 1985-1987 to 2018-2020 | -0.0161 | -0.0061 | -0.0033 | -0.0035 | -0.0059 | -0.0085 | -0.0097 | -0.0530 |
| Contribution of change in marital fertility |  |  |  |  |  |  |  |  |
| 1985-1987 to 1991-1993 | -0.0005 | -0.0003 | -0.0013 | -0.0023 | -0.0023 | -0.0027 | -0.0033 | -0.0126 |
| 1991-1993 to 1994-1996 | -0.0011 | -0.0001 | -0.0002 | 0.0001 | -0.0011 | -0.0006 | -0.0031 | -0.0061 |
| 1994-1996 to 2004-2006 | 0.0006 | -0.0001 | -0.0009 | -0.0025 | -0.0036 | -0.0044 | -0.0016 | -0.0125 |
| 2004-2006 to 2011-2013 | 0.0006 | 0.0004 | -0.0002 | -0.0013 | -0.0023 | -0.0039 | -0.0060 | -0.0127 |
| 2011-2013 to 2016-2018 | 0.0006 | -0.0002 | 0.0005 | 0.0015 | 0.0014 | 0.0015 | 0.0022 | 0.0075 |
| 2016-2018 to 2018-2020 | 0.0006 | 0.0002 | -0.0001 | 0.0001 | 0.0005 | 0.0004 | 0.0006 | 0.0023 |
| 1985-1987 to 2018-2020 | 0.0022 | 0.0001 | -0.0015 | -0.0032 | -0.0060 | -0.0089 | -0.0099 | -0.0272 |
| Contribution of change in proportion of women married |  |  |  |  |  |  |  |  |
| 1985-1987 to 1991-1993 | -0.0017 | -0.0006 | -0.0002 | 0.0000 | 0.0000 | 0.0001 | -0.0005 | -0.0028 |
| 1991-1993 to 1994-1996 | -0.0009 | 0.0003 | 0.0002 | 0.0001 | 0.0000 | 0.0001 | 0.0002 | -0.0001 |
| 1994-1996 to 2004-2006 | -0.0024 | -0.0006 | -0.0002 | -0.0001 | -0.0001 | 0.0000 | -0.0001 | -0.0034 |
| 2004-2006 to 2011-2013 | -0.0031 | -0.0010 | -0.0003 | 0.0000 | 0.0000 | 0.0000 | 0.0001 | -0.0042 |
| 2011-2013 to 2016-2018 | -0.0048 | -0.0015 | -0.0004 | -0.0001 | 0.0000 | 0.0001 | 0.0001 | -0.0065 |
| 2016-2018 to 2018-2020 | -0.0008 | -0.0008 | -0.0003 | -0.0001 | 0.0000 | 0.0000 | 0.0001 | -0.0019 |
| 1985-1987 to 2018-2020 | -0.0183 | -0.0061 | -0.0018 | -0.0003 | 0.0001 | 0.0004 | 0.0003 | -0.0258 |

Source: Author

Tables 5 and 6 also reveal that the contribution of the change in age-specific marital fertility rates to the change in the arithmetic mean of age-specific fertility rates (*f*_*a*_) and in the geometric mean of age-specific fertility rates (*f*_*g*_) has been different in different time-segments of the period 1985-2020. During the period 2011-2020, the change in age-specific marital fertility rates contributed to the increase, instead decrease in both *f*_*a*_ and *f*_*g*_ as the age-specific marital fertility rates increased during this period. On the other hand, change in the age-specific proportion of married women contributed to the decrease in both *f*_*a*_ and *f*_*g*_. However, the contribution of the change in the age-specific marital fertility rates to the change in *f*_*a*_ is offset by the contribution of the change in the age-specific proportion of married women so that *f*_*a*_ decreased during this period. On the other hand, the contribution of the change in the age-specific proportion of married women to the change in *f*_*g*_ could not offset the contribution of the change in age-specific marital fertility rate so that *f*_*g*_ increased during the period.

Table 5 also suggests that the main contributor to the decrease in *f*_*a*_ during the period 1985-2020 has been the decrease in fertility in the age group 20-24 years (33 per cent) whereas the decrease in fertility in the age group 15-19 years accounted for a decrease of almost 19 per cent and the decrease in fertility in the age group 25-29 years accounted for a decrease of almost 18 per cent. This means that almost 70 per cent of the decrease in *f*_*a*_ is due to the decrease in fertility in the age-group 15-29 years. The contribution of change in marital fertility and in the proportion of married women in different age groups to the change in fa. has, however, been different. Fertility in the age-group 15-24 years decreased despite the increase in marital fertility in this age group because of the decrease in the proportion of married women aged 15-24 years.

Table 6 suggests that the decrease in fertility in the age-group 15-29 years has accounted for less than 50 per cent of the decrease in the geometric mean of age-specific fertility rates, *f*_*g*_, during this period while the decrease in fertility in the age-group 30 years and above has accounted for slightly more than 50 per cent of the decrease in *f*_*g*_. The geometric mean of age-specific fertility rates, *f*_*g*_, increased during the period 2011-2020 mainly because of the increase in fertility in the age group 30-49 years. It is obvious from tables 5 and 6 that the contribution of the change in age-specific marital fertility rates and the change in the age-specific proportion of married women to the change in the fertility experience of women of childbearing ages is different when the simple arithmetic mean is used as the aggregation function and when the geometric mean is used as the aggregation function to aggregate the age-specific fertility rates. The sensitiveness of the change in fertility to the aggregation function used to aggregate age-specific fertility rates bears importance in the analysis of fertility change.

### Transition in Age Pattern of Fertility/Marital Fertility

It is well-known that fertility by age because both marital fertility and proportion of married women vary by age. Since fertility is not the same in all ages of the childbearing period, the simple arithmetic mean of age-specific fertility rates (*f*_*a*_) is not equal to the geometric mean of the age-specific fertility rates (*f*_*g*_). The simple arithmetic mean of the age-specific fertility rates is equal to the geometric mean of age-specific fertility rates only when fertility rates different ages of the childbearing period are the same. Otherwise, the simple arithmetic mean of age-specific fertility rates is always greater than the geometric mean of age-specific fertility rates. The ratio *f*_*a*_/*f*_*g*_, therefore, reflects the variation or the inequality in fertility in different ages of the childbearing period. It is also obvious that the larger this ratio from the limiting value of 1, the larger the variation or inequality in fertility in different ages of the childbearing period. By the same argument, the ratio *g*_*a*_/*g*_*g*_ reflects the variation or the inequality in marital fertility in different ages of the childbearing period. The trend in the ratio *f*_*a*_/*f*_*g*_, therefore, reflects the change in the age pattern of fertility with the change in fertility. Similarly, the change in the ratio ga/gg reflects the change in the age pattern of marital fertility with the change in marital fertility. The change in the age pattern of fertility has implications for future population growth and population stabilisation as concentration of fertility in younger ages has implications for future population growth in terms of population momentum.

Figure 6 shows the trend in the ratios *f*_*a*_/*f*_*g*_ and *g*_*a*_/*g*_*g*_ in India during 1985-2020. Both the ratios increased during the period 1985-2014 which suggests that there has been an increase in the concentration of both fertility and marital fertility in selected ages of the childbearing period with the decrease in fertility and marital fertility during this period. The inequality in fertility or marital fertility, however, decreased after 2012-2014. The decrease in the age inequality of marital fertility has been associated with the increase in marital fertility. On the other hand, the decrease in the inequality in fertility by age has been associated with the marginal increase in *f*_*g*_ but a decrease in *f*_*a*_. From the perspective of population stabilisation, it is important that the decrease in fertility should be associated with the increase in dispersion of fertility across childbearing ages as it contributes to lower the impact of momentum on population growth.

**Figure 6:**
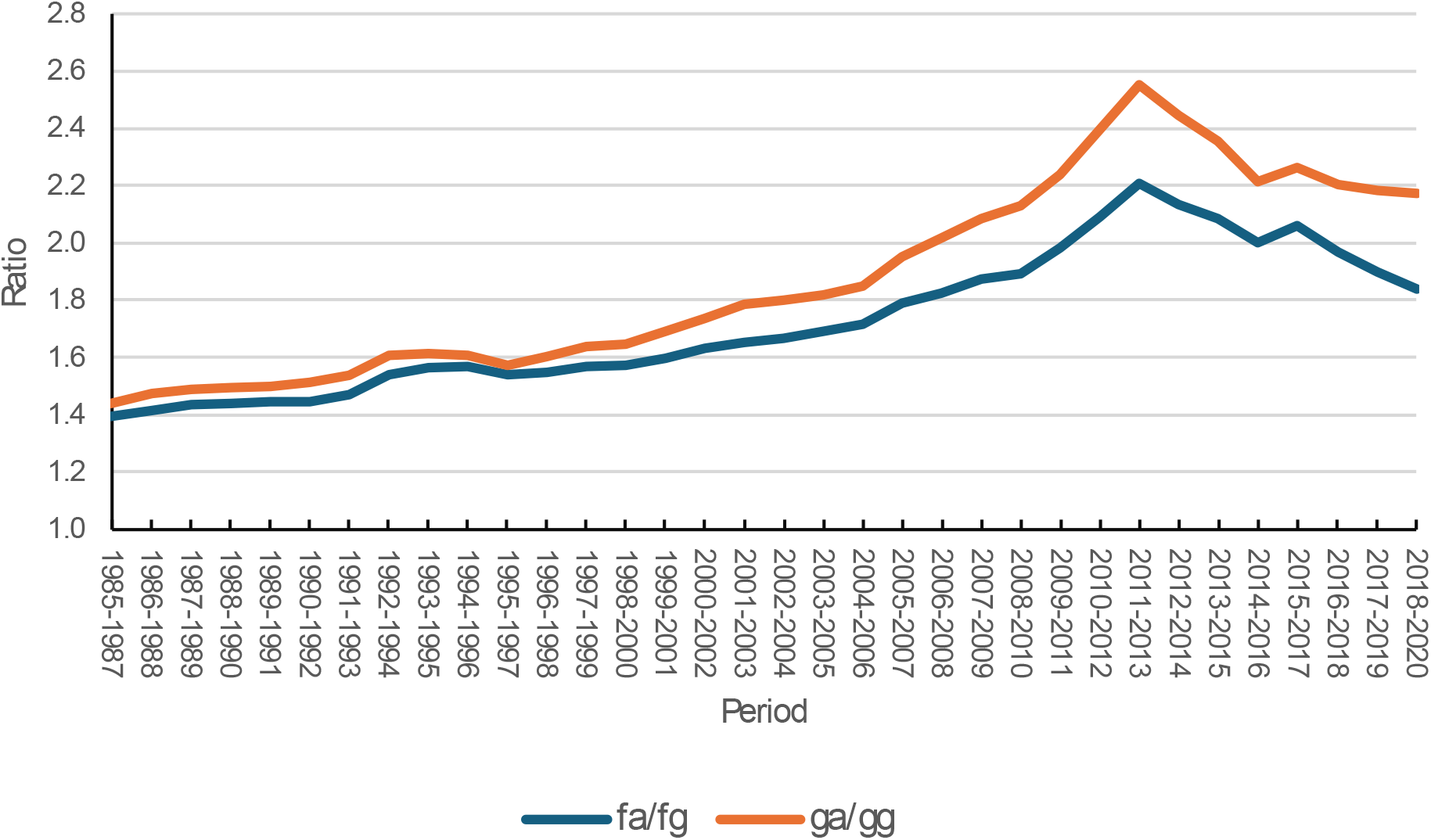
Trend in age inequality of fertility (*f*_*a*_/*f*_*g*_) and age inequality of marital fertility (*g*_*a*_/*g*_*g*_) in India, 1985-2020. Source: Author

The trend in the inequality in fertility and marital fertility by age is supported by the trend in the mean age of childbearing of women (MACB_w_) and the mean age of childbearing of married women (MACB_m_). The decreasing trend in both MACB_w_ and MACB_m_ during 1985-2013 confirms increasing concentration of both fertility and marital fertility in the younger ages of the childbearing period. After 2011-2013, the MACB_w_ increased rather sharply but the increase in MACB_m_ has been relatively modest which indicates a marked shift in the age pattern of fertility, but only a marginal shift in the age pattern of marital fertility (Figure 7). The joinpoint regression analysis reveals that average annual per cent change (AAPC) in MACB_w_ was 0.084 per cent during 1985-2020 which indicates that there has been only a marginal shift in the age pattern of fertility towards older ages of the childbearing period. The MACB_w_ increased during the time-segments 1992-2000 and 2012-2020 only. On the other hand, the average annual per cent change (AAPC) in the mean age of fertility schedule of married women of childbearing age (MACB_m_) was -0.283 per cent which indicates that fertility of married women has got increasingly concentrated in the younger ages of the childbearing period with the decrease in marital fertility, although MACBm increased during the time-segments 1992-1997 and 2011-2016. The MACB_m_ during 2018-2020 was also markedly lower than MACB_m_ during 1985-1987, while MACB_w_ during 2018-2020 was only marginally higher than MACB_w_ during 1985-1987. It appears that there has been little change in the age pattern of fertility and in the age pattern of marital fertility in the country with the decrease in fertility and marital fertility. This means that the change in fertility in India during 1985-2020 has contributed virtually little to lower the impact of momentum on the future population growth and hence to population stabilization.

**Figure 7:**
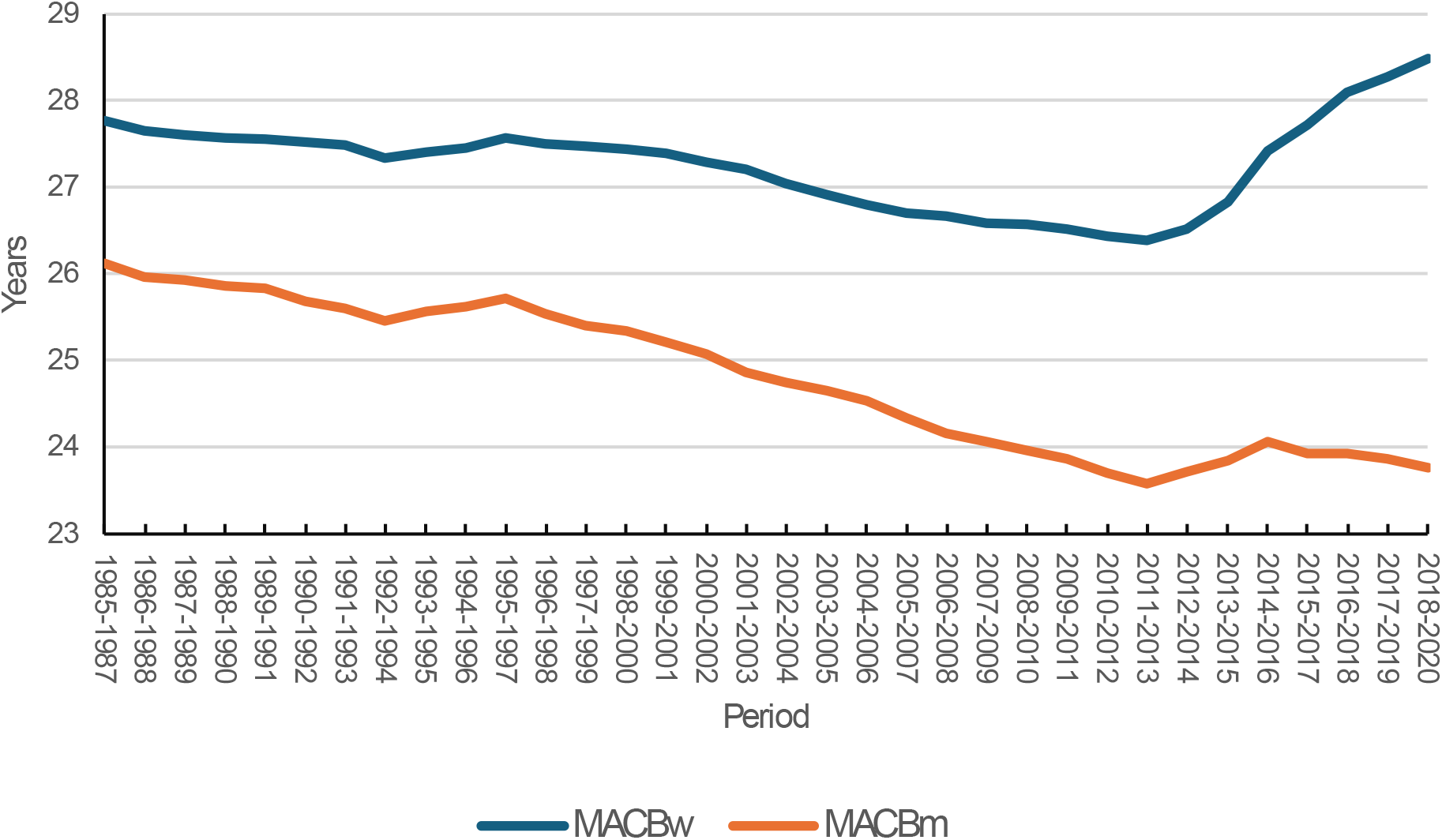
Mean age of childbearing in women of childbearing age and in married women of childbearing age in India, 1985-2020. Source: Author

## Discussions and Conclusions

The present paper reveals two different perspectives of fertility transition in India depending upon the way the fertility experience of women of childbearing age is aggregated. The simple arithmetic mean of age-specific fertility rates suggest that fertility in India has decreased consistently throughout the period 1985-2020, although at varying pace. The geometric mean of age-specific fertility rates, however, suggests that the decrease in fertility has stalled during 2011-2020. The reason is that the decrease in fertility has been different in different ages of the childbearing period. Fertility in the age group 15-29 years has decreased throughout the period 1985-2020 but fertility in the age group 30-49 years appears to have increased after 2011-2013. The decrease in fertility ages 15-29 years has, however, been more than the increase in fertility in ages 30-49 years so that simple arithmetic mean of age-specific fertility rates and hence TFR decreased during 2011-2020. The trend in the simple arithmetic mean of age-specific fertility rates hides the increase in fertility in older ages during this period. This is not the case with the geometric mean. The change in geometric mean weights equally the change in fertility in different ages whereas the change in simple arithmetic mean weights the change in fertility in different ages in proportion to the size of the change (2021). From the perspective of the transition in the fertility experience of women of childbearing age, it is more appropriate to aggregate age-specific fertility rates using the geometric mean rather than using simple arithmetic mean as the aggregation function.

The selection of the function to aggregate the age-specific fertility rates also influences the contribution of the change in the age-specific marital fertility rates and the change in the proportion of married women in different ages to the change in the composite measure of fertility. When the geometric mean is used as the aggregation function, the change in marital fertility and the change in the proportion of married women contributes almost equally to the change in the geometric mean of the age-specific fertility rates. However, when the simple arithmetic mean is used to aggregate age-specific fertility rates, the contribution of the change in marital fertility is substantially lower than the contribution of the change in the proportion of married women leading to the conclusion that transition has been driven primarily by the change in the proportion of married women. The TFR in India has been associated with the decrease in TMFR during 1985-2013 but the decrease in TFR during 2011-2020 has been associated with the increase in TMFR which implies that the entire decrease in TFR during the post 2011 period has been due to the decrease in the proportion of married women. This conclusion appears untenable as marriage is nearly universal in India and female marriage at an early age is quite common despite the fact that marriage of a girl younger than 18 years of age is not legally permitted. In contrast, the geometric mean of age-specific fertility rates indicates that stalling of fertility transition in the post 2011 period has been due to the increase in marital fertility in older ages of the childbearing period.

Another unique feature of fertility transition in India is that the decrease in fertility has been associated with the creased concentration of fertility in selected ages of the childbearing period as is reflected through the increase in the ratio of simple arithmetic mean to the geometric mean of age-specific fertility rates and the decrease in the mean age at childbearing. This trend reflects the typical approach adopted by India to regulate fertility of married women which has always focused on birth limitation rather than birth spacing. The ratio of the simple arithmetic mean to the geometric mean of the age-specific birth rates and the mean age at childbearing increased after 2011-2013 because of the increase in the fertility of older women.

The present analysis suggests that fertility transition based on the simple arithmetic mean of age-specific fertility rates or TFR should be interpreted with caution as the change in TFR hides more than what it reveals and the change in the fertility behaviour of women of childbearing age. Rather any analysis of fertility transition should be based on the geometric mean of age-specific fertility rates as the change in geometric mean assigns equal weight to the change in fertility in different ages of the childbearing period.

The present analysis highlights some concerns related to fertility transition in India that have policy and programme implications. There is a need to look into the reasons behind the increase in fertility of older women, especially after 2011-2013. There may be a possibility that the decrease in fertility of older married women is small because of low fertility of these women and this small decrease is offset by the increase in fertility because of the increase in the proportion of married women because of the improvement in mortality in the older ages of the childbearing period. Another possible factor may be the shift in the focus of official family planning efforts to spacing methods of contraception in place of terminal methods of contraception. The increase in fertility of married women 2011-2013 needs to be looked in terms of its three proximate determinants breastfeeding behaviour, family planning use, and practice of abortion (Bongaarts, 1978; Preston et al, 2001), The present analysis suggests that the increase in marital fertility appears to be the reason behind stalling of fertility transition after 2011.

## Data Availability

All data used are data published by the Registrar General of India.

## References

Bhatia R (2008) The logarithmic mean. Resonance June 2008: 583–594.

Bongaarts J (1978) A framework for analyzing the proximate determinants of fertility. Population and Development Review 4(1): 105–132.

Bongaarts J (1994) Population policy options in the developing world. New York, Population Council, Policy Research Division Working Paper No. 59.

Bongaarts J, Feeney G (1998) On the quantum and tempo of fertility. Population and Development Review 24(2): 271–91.

Bullen PS (2000) Handbook of Means and Their Inequalities. Dordrecht, Netherlands, Kluwer: 175–177.

Carlson BC (1966). “Some inequalities for hypergeometric functions. Proceedings of American Mathematical Society 17: 32–39.

Desai MJ (1991) Human development concepts and measures. European Economic Review 35(2-3): 350–357.

Feeney G (1983) Population dynamics based on birth intervals and parity progression. Population Studies 37: 77–89.

Frejka T (1982) Momentum. In JA Ross (ed) International Encyclopedia of Population 2: 450–451. New York, Free Press.

Government of India (1969) The Registration of Births and Deaths Act 1969. New Delhi, Ministry of Home Affairs.

Government of India (1988) Census of India 1981. Fertility in India. An Analysis of 1981 Census Data. New Ministry of Home Affairs, Office of the Registrar General, India. Demography Division.

Government of India (1997) District Level Estimates of Fertility and Child Mortality for 1991 and their Relations with Other Variables. New Delhi, Registrar General India. Occasional Paper 1 of 1997.

Government of India (2000) National Population Policy 2000. New Delhi, Ministry of Health and Family Welfare.

Government of India (2022a) Sample Registration System Statistical Report 2022. New Delhi, Ministry of Home Affairs, Office of the Registrar General and Census Commissioner of India.

Government of India (2022b) National Family Health Survey (NFHS-5) India Report. New Delhi, Ministry of Health and Family Welfare.

Government of India (2023) The Registration of Births and Deaths (Amendment) Act 2023. New Delhi, Ministry of Law and Justice.

Guilmoto CZ, Rajan I (2002) District level estimates of fertility from India’s 2001 Census. Economic and Political Weekly 37(7): 665–672.

Guilmoto CZ, Rajan I (2011) Fertility at district level in India: Lessons from the 2011 Census. Paris, Université Paris Descartes, INED, IRD. Working Paper du CEPED, n°30.

Hajnal J (1947) The analysis of birth statistics in the light of the recent international recovery of the birth-rate. Population Studies 1: 137–64.

Keyfitz N (1971) On the momentum of population growth. Demography 8(1): 71–80.

Kim H-J, Fay MP, Feuer EJ, Midthune DN (2000) Permutation tests for joinpoint regression with applications to cancer rates. Statistics in Medicine 19: 335–351.

Kim H-J, Yu B, Feuer EJ (2009) Selecting the number of change-points in segmented line regression. Statistica Sinica 19(2): 597–609.

Kim H-J, Chen H-S, Midthune D, Wheeler B, Buckman DW, Green D, Byrne J, Luo J, Feuer EJ (2022) Data-driven choice of a model selection method in Joinpoint Regression. Journal of Applied Statistics 1–22.

Kim J, Kim H-J (2016) Consistent model selection in segmented line regression. Journal of Statistical Planning and Inference 170:106–116.

Klugman J, Rodriguez F, Choi H-J (2011) The HDI 2010: new controversies, old critiques. New York, United Nations Development Program. Human Development Research Paper, 2011/01.

Kovacevic M (2010) Review of HDI critiques and potential improvements. New York, United Nations Development Programme. Human Development Research Paper No. 2010/33.

Lermon PM (1980) Fitting segmented regression models by grid search. Journal of Royal Statistical Society, Series C (Applied Statistics), 29(1), 77–84.

Matthews GE (2021) When averageing multiples, the arithmetic mean is inferior to the harmonic mean. Business Valuation Review 61–67.

Merrick TW (1986) World population in transition. Population Bulletin 41(2): 1–52.

Marriot LD (2010) Colorectal cancer network (CRCNet) user documentation for surveillance analytic software: Joinpoint. Cancer Care Ontario: 1–28.

Mishra VK, Palmore JA, Sinha SK (1994) Indirect estimates of fertility and mortality at the district level, 1981. New Delhi, Office of the Registrar General, India.

National Cancer Institute (2023) Joinpoint Regression Program, Version 5.0.2. Statistical Methodology and Applications Branch, Surveillance Research Program, National Cancer Institute.

Park N, Vyas S, Broussard K, Spears D (2023) Near-universal marriage, early childbearing, and low fertility: India’s alternative fertility transition. Demographic Research 48(34): 945–956.

Preston SH, Heuveline P, Guillot M (2001) Demography. Measuring and Modeling Population Processes. Oxford, Blackwell Publishing.

Ryder NB (1964) The process of demographic translation. Demography 1: 74–82.

United Nations (2022) World Population Prospects 2022. Online Edition. New York, Department of Economic and Social Population Division.

United Nations (2024) World Population Prospects 2024. Online Edition. New York, Department of Economic and Social Population Division.

Zhang NR, Siegmund DO (2007) A modified Bayes Information Criterion with applications to the analysis of comparative genomic hybridization data. Biometrics 63(1): 22–32.

